# Cognitive reserve in ALS: The role of occupational skills and requirements

**DOI:** 10.1101/2023.06.21.23291677

**Authors:** Emma Rhodes, Sebleh Alfa, Hannah Jin, Lauren Massimo, Lauren Elman, Defne Amado, Michael Baer, Colin Quinn, Corey T. McMillan

## Abstract

**Background:** Amyotrophic Lateral Sclerosis (ALS) is a heterogeneous neurodegenerative condition featuring variable degrees of motor decline and cognitive impairment. We test the hypothesis that cognitive reserve (CR), defined by occupational histories involving more complex cognitive demands, may protect against cognitive decline, while motor reserve (MR), defined by working jobs requiring complex motor skills, may protect against motor dysfunction.

**Methods:** Individuals with ALS (n=150) were recruited from the University of Pennsylvania’s Comprehensive ALS Clinic. Cognitive performance was evaluated using the Edinburgh Cognitive and Behavioral ALS Screen (ECAS), and motor functioning was measured using Penn Upper Motor Neuron (PUMNS) scale and ALS Functional Rating Scales (ALSFRS-R). The Occupational Information Network (O*NET) Database was used to derive 17 factors representing distinct worker characteristics, occupational requirements, and worker requirements, which were related to ECAS, PUMNS, and ALSFRS-R scores using multiple linear regression.

**Results:** A history of working jobs involving greater reasoning ability (β=2.12, p<.05), social ability (β=1.73, p<.05), analytic skills, (β=3.12, p<.01) and humanities knowledge (β=1.83, p<.01) was associated with better performance on the ECAS, while jobs involving more exposure to environmental hazards (β=-2.57, p<.01) and technical skills (β=-2.16, p<.01) were associated with lower ECAS Total Scores. Jobs involving greater precision skills (β=1.91, p<.05) were associated with greater disease severity on the PUMNS. Findings for the ALSFRS-R did not survive correction for multiple comparisons.

**Discussion:** Jobs requiring greater reasoning abilities, social skills, and humanities knowledge were related to preserved cognitive functioning consistent with CR, while jobs with greater exposure to environmental hazards and technical demands were linked to poorer cognitive functioning. We did not find evidence of MR as no protective effects of occupational skills and requirements were found for motor symptoms, and jobs involving greater precision skills and reasoning abilities were associated with worse motor functioning. Occupational history provides insight into protective and risk factors for variable degrees of cognitive and motor dysfunction in ALS.

## Introduction

Amyotrophic Lateral Sclerosis (ALS) is a devastating multi-system disorder characterized by motor neuron degeneration. Originally thought to solely affect motor functioning, there is increasing evidence that up to 40% of individuals with ALS will develop cognitive impairment in at least one domain^1^ with selective deficits in verbal fluency and other expressive language skills, executive functioning, and social cognition.^2^ While multiple risk factors for cognitive impairment in ALS have been identified, including the presence of *C9orf72* repeat expansions, family history of ALS, bulbar symptom onset, and predominant upper motor neuron (PUMN) phenotype,^3^ less is known about resilience factors that may protect against cognitive impairment. Emerging work has identified a protective effect of educational and occupational attainment on cognitive functioning in individuals with ALS, highlighting a novel role for the cognitive reserve (CR) hypothesis in motor neuron disease^4–8^ but existing research has used gross metrics of occupational and educational attainment that may obscure the impact of specific skills and traits on cognitive functioning. Furthermore, it remains unclear if CR contributes to attenuated decline in motor functioning in ALS.

The CR hypothesis posits that enriched life experiences, including educational attainment, occupational complexity, and physical activity, contribute to preserved cognitive functioning in the face of brain injury or illness.^9^ The protective effect of CR has been observed in multiple neurodegenerative diseases, including Alzheimer’s disease,^10^ frontotemporal dementia,^7^ and Parkinson’s disease.^11^ CR is typically measured using proxy variables that reflect high premorbid functioning and enriched life experiences, such as education level, occupational attainment, premorbid IQ, and frequency of exercise and other leisure activities.^12^ Studies of CR in ALS have used various individual proxies and composite scores that combine multiple CR proxies and demographic features, including education and premorbid IQ;^5^ years of education,^4, 7^ educational and occupational attainment, midlife leisure activities, and bilingualism;^8^ and verbal intelligence.^5, 6^ Cross-sectional evidence has found these CR proxies to be associated with better performance on tests of global cognition, verbal fluency, working memory, verbal learning and memory, and visuospatial abilities in ALS.^6, 8^ Longitudinal work has been more limited and equivocal, with one study reporting attenuated decline in verbal fluency among ALS patients with high CR^6^ and another showing that CR’s protective effect was limited to the level of cognitive performance at baseline with no measurable effect on the slope of cognitive change over time.^13^

Occupational attainment is a robust proxy for CR that shows moderate to strong associations with preserved cognition across multiple clinical populations,^9, 14, 15^ including ALS.^8, 13^ High levels of occupational attainment are thought to promote CR via multiple pathways, including links to higher socioeconomic status and better access to healthcare and stable housing, which can mitigate medical risk throughout the lifespan, as well as the “use it or lose it” hypothesis, in which long term engagement in complex mental activities stimulates neural activation that buffers against decline in cognition. Evidence for the “use it or lose it” hypothesis can be found in studies of CR in Alzheimer’s disease and related dementias showing that the protective effect of high educational attainment disappears after accounting for occupational complexity,^16, 17^ suggesting that continued use of cognitive skills is needed to foster CR. Further, occupational complexity has demonstrated stronger associations with gray matter integrity relative to education.^15^ Taken together, occupational complexity and attainment have been highlighted as salient and robust CR proxies in ALS and other neurodegenerative diseases, but less is known about how specific occupational skills and requirements may impart reserve for individuals at risk for cognitive impairment, which could help clarify mechanisms of CR and identify occupations with the most potential to have a protective effect.

In addition to cognitive skills, many occupations also involve specific motor activities and abilities that could moderate expression of motor dysfunction in ALS. Motor reserve is an emerging concept with preliminary support from PD research showing that high CR is associated with reduced motor dysfunction in early stages of disease, but not rate of decline.^11^ Evidence for motor reserve in ALS is more limited, but one study found a protective effect of a CR composite that included occupation on motor functioning, but only in individuals with bulbar symptom onset.^8^ Other groups have identified strenuous, repetitive physical activity as a risk factor for ALS,^18–20^ but it remains unclear if occupational factors contribute to this effect.

The goal of this study was to assess the impact of specific occupational skills and requirements on cognitive and motor functioning in ALS. We used empirically derived factor scores to represent categories of common job skills and requirements and tested their associations with validated measures of cognitive and motor functioning in ALS. We hypothesized that a history of working occupations high on factors representing more complex or executively demanding job skills would be associated with better cognitive performance in domains typically impacted by ALS (i.e., verbal fluency, executive functioning, social cognition). Additionally, we predicted that an occupational history high in factors reflecting greater motor skills and demands would be associated with worse motor functioning.

## Methods

### Participants

Participants included 150 individuals with ALS who were recruited for research at the University of Pennsylvania (UPenn) Comprehensive ALS Clinic and Frontotemporal Degeneration Center. Inclusion criteria were a clinical diagnosis of probable or definite ALS made by a board-certified neurologist using the El Escorial criteria,^21^ and ability to complete cognitive testing. Exclusion criteria included the presence of other neurologic (e.g., stroke, epilepsy) or psychiatric (e.g., schizophrenia, major depressive disorder) conditions that would compromise performance on cognitive testing. Six participants met criteria for comorbid frontotemporal dementia (FTD), which was not considered exclusionary based on considerable clinical and etiologic overlap with ALS.^22^

### Procedures

All participants completed a comprehensive clinical evaluation including physical and neurologic exams and cognitive testing within one year of ALS diagnosis as part of ongoing observational research. Site (e.g., limb, bulbar) and date of motor symptom onset were recorded for each participant. This project was conducted in accordance with the Helsinki Declaration and was approved by the UPenn Institutional Review Board.

### Measures

#### Edinburgh Cognitive and Behavioral Assessment Screen (ECAS)

The ECAS is a brief neuropsychological assessment tool designed specifically for ALS to accommodate physical challenges (e.g., limited ability to write or speak) that can confound traditional neuropsychological testing in this population.^23^ The ECAS measures performance in five cognitive domains: language, fluency, executive function, memory, and visuospatial function. This study used the North American ECAS,^2^ which is modified to include culturally appropriate stimulus items distinct from the original UK forms (e.g., shopping trolleys ➔ shopping carts). Primary outcome variables from the ECAS include raw scores for each ECAS cognitive domain and composite scores for ALS Specific (i.e., language, executive function, verbal fluency) and ALS Non-Specific (i.e., memory and visuospatial function) abilities. Composite scores are computed as the sum of raw scores from the relevant domains.

#### Revised ALS Functional Rating Scale (ALSFRS-R)

The ALSFRS-R is a 10-item Likert scale used to evaluate the disease severity and functional status of patients with ALS.^24^ It measures the presence and severity of everyday problems with speech, salivation, swallowing, handwriting, cutting food and handling utensils, dressing and hygiene, turning in bed and adjusting bed clothes, walking, climbing stairs, and breathing on a 0 (no function) to 4 (normal function) scale. The maximum score is 48, and a higher score reflects better functioning.

#### Penn Upper Motor Neuron Score (PUMNS)

The PUMNS is a 28-item scale used to assess clinical upper motor neuron impairment in ALS.^25^ It includes separate subscores for bulbar (range: 0-4), upper extremity (range: 0-7), and lower extremity (range: 0-7) with a total possible score of 32. Higher scores indicate greater upper motor neuron disease burden. The PUMNS has high inter-rater reliability (all coefficients >0.96) and internal validity with subscore correlation coefficients ranging from 0.68 to 0.85.

#### Occupational History

Data on participant occupational history was obtained as part of the clinical evaluation. For individuals with a history of more than one occupation, the highest or most advanced level was selected for analysis. Participants’occupations were categorized using the version 22.3 of the Occupational Information Network (O*NET) Database, which includes data on hundreds of standardized job descriptors based on the Standard Occupation Classification (O*NET-SOC) taxonomy. O*NET descriptors are categorized into six overlapping domains: Occupational Requirements, Worker Characteristics, Worker Requirements, Occupation-Specific Information, Experience Requirements, and Workforce Characteristics.^26^ We used variables from Occupational Requirements, Worker Characteristics, and Worker Requirements domains as descriptors, as these domains describe broad features of occupational skills and requirements and are commonly used in other studies seeking to relate O*NET descriptors to health outcomes.^27^ Worker Characteristics are enduring characteristics and abilities of individuals (e.g., memorization) influencing work performance, while Worker Requirements refer to work-related knowledge and skill (e.g., mathematics). Occupation Requirements include work activites (e.g., getting information) and physical and social contexts (e.g., contact with others). O*NET data for Worker Characteristics, Worker Requirements, and Occupational Requirements were retrieved for each participant’s reported highest level of occupation. The Occupational Requirements (98 descriptors), Worker Characteristics (80 descriptors), Worker Requirements (68 descriptors) domains include 246 descriptors in total. We rescaled all descriptors so that they ranged from 0 to 100. We then performed principal component analysis (PCA) within each of the three domains, using 966 standardized O*NET occupations as observations. Factors extracted from the PCA were used as independent variables in separate linear regression models evaluating the impact of occupation-specific Worker Characteristics, Worker Requirements, and Occupational Requirements on cognitive and motor functioning in our ALS sample (see Statistical Analyses).

### Statistical Analyses

Dimensionality of O*NET Worker Characteristics, Worker Requirements, and Occupational Requirements descriptors was reduced using PCA, because the O*NET model does not define clear relationships among the descriptors within its various categories and domains.^28^ To determine the number of components to extract, we retained the first several components that together explained about 70% of the variance in the data within each domain and examined scree plots. Seven components explained 71.0% of the variance within the Occupational Requirements domain. Five components explained 73.0% of the variance within the Worker Characteristics domain. Five components explained 70.8% of the variance within the Worker Requirements domain. We then performed a varimax rotation within each of the three Domains to maximize the variance of the squared loadings within each component. ^29, 30^ In total, we retained 17 components from across the three Domains (Supplemental Tables 1-3).

Continuous variables were summarized with mean and SD. Categorical variables were summarized with percentages. Separate multiple linear regression models were used to assess associations between components derived for Occupational Requirements, Worker Characteristics, Worker Requirements, and cognitive and motor functioning covarying for age, education, and bulbar symptom onset. Outcome variables were examined for normality using histograms, Shapiro-Wilk tests, and Q-Q plots. Models were assessed for collinearty using variance inflation factor tests (all values > 10) and heteroskedasticity using scale-location plots and Breusch-Pagan tests (p > .05 for all models). Cook’s distance > 4/n was used to identify outliers, resulting in 8 observations being removed from models predicting cognitive functioning (Table 3). Over 2/3 of our sample (n = 103) had genetic testing results for C9orf72 repeat expansions, and a separate set of regression analyses was performed in this subset as described above but additionally covarying for C9orf72 status (Supplemental Tables 4-5). Omnibus regression models were corrected for multiple comparisons using Bonferroni correction with a cutoff of *p* < .005.

## Results

### Sample Characteristics

The sample consisted of 150 individuals with a clinical diagnosis of ALS (Table 1). The mean age was 62.66 years (SD = 10.02) and mean years of education was 14.95 (SD = 2.64). Mean disease duration was 2.93 years (SD = 2.9), and mean diagnostic delay (i.e., time from symptom onset to diagnosis) was 1.48 years (SD = 1.6). Ninety-two percent of the sample identified as white (n = 138), and 98% identified as non-Hispanic (n = 147). The mean ECAS Total score was 107.64 (SD = 13.06), and 85.33% of the sample had ECAS total scores in the unimpaired range using demographically adjusted normative correction.^2^ The mean PUMNS Total score was 9.01 (SD = 8.18), and the mean FRS Total score was 33.49 (SD = 7.12). Seventy-four percent (n = 111) of the sample had non-bulbar onset of ALS symptoms, 22.67% (n = 34) had bulbar onset, and onset site was unknown for 3.33% (n = 5). A subset of the larger sample (n = 103) had genotyping for C9orf72 repeat expansions, with 61.3% (n = 92) C9 negative and 7.3% (n = 11) C9 positive. The majority of the sample reported working white collar jobs (n = 111, 74%), such as manager, administrator, or clerical or professional worker, while 26% of the sample (n = 39) reported working blue collar jobs, such as service worker, craftsman, foreman, or laborer (Figure 1).

**Table 1.**
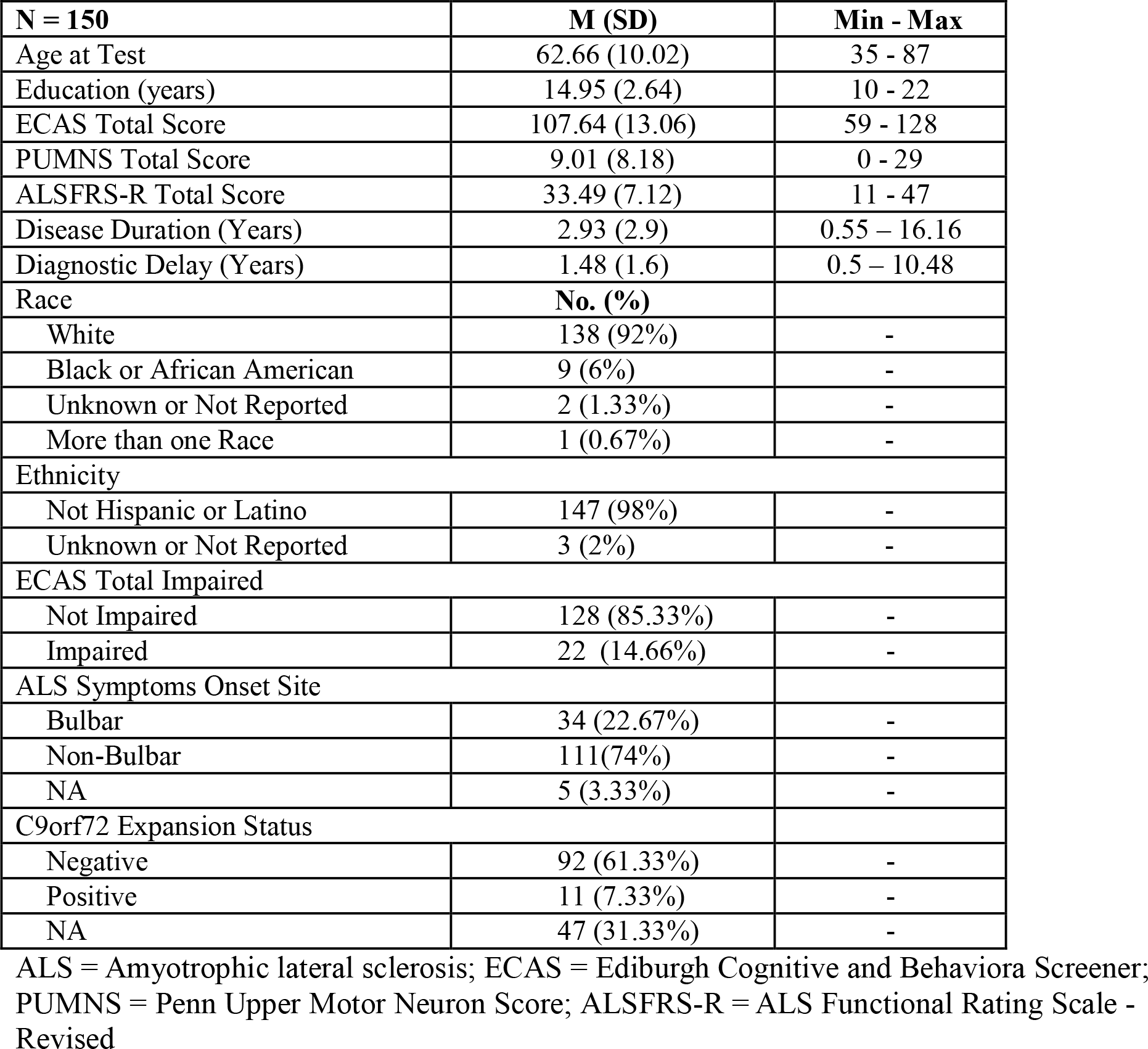
Sample Characteristics

| **N = 150** | **M (SD)** | **Min - Max** |
| --- | --- | --- |
| Age at Test | 62.66 (10.02) | 35 - 87 |
| Education (years) | 14.95 (2.64) | 10 - 22 |
| ECAS Total Score | 107.64 (13.06) | 59 - 128 |
| PUMNS Total Score | 9.01 (8.18) | 0 - 29 |
| ALSFRS-R Total Score | 33.49 (7.12) | 11 - 47 |
| Disease Duration (Years) | 2.93 (2.9) | 0.55 – 16.16 |
| Diagnostic Delay (Years) | 1.48 (1.6) | 0.5 – 10.48 |
| Race | **No. (%)** |  |
| White | 138 (92%) | - |
| Black or African American | 9 (6%) | - |
| Unknown or Not Reported | 2 (1.33%) | - |
| More than one Race | 1 (0.67%) | - |
| Ethnicity |  |  |
| Not Hispanic or Latino | 147 (98%) | - |
| Unknown or Not Reported | 3 (2%) | - |
| ECAS Total Impaired |  |  |
| Not Impaired | 128 (85.33%) | - |
| Impaired | 22 (14.66%) | - |
| ALS Symptoms Onset Site |  |  |
| Bulbar | 34 (22.67%) | - |
| Non-Bulbar | 111(74%) | - |
| NA | 5 (3.33%) | - |
| C9orf72 Expansion Status |  |  |
| Negative | 92 (61.33%) | - |
| Positive | 11 (7.33%) | - |
| NA | 47 (31.33%) | - |

ALS = Amyotrophic lateral sclerosis; ECAS = Ediburgh Cognitive and Behaviora Screener; PUMNS = Penn Upper Motor Neuron Score; ALSFRS-R = ALS Functional Rating Scale - Revised

**Table 2.** Multiple linear regression models of cognitive performance with O*NET Occupation Requirements (A), Worker Characteristics (B), and Worker Requirements (C)

| **A. Occupational Requirements** | **ECAS Total Score** | **ECAS ALS Specific Score** | **ECAS ALS Non-Specific Score** |
| --- | --- | --- | --- |
| Model R<sup>2</sup> | 0.31 | 0.28 | 0.08 |
| F-statistic | 7.17 | 6.35 | 3.11 |
| DF | 11, 118 | 11, 118 | 11, 118 |
| *p* | 1.30e-09 | 1.675e-07 | .003 |
|  | β (S.E.) | β (S.E.) | β (S.E.) |
| Intercept | 112.97 (7.13)*** | 85.29 (6.18)*** | 28.23 (2.84)*** |
| Age at Test | -0.33 (0.08)*** | -0.26 (0.07)*** | -0.07 (0.03)* |
| Education | 1.10 (0.35)** | 0.83 (0.31)** | 0.31 (0.14)* |
| Non-Bulbar Onset | -0.75 (1.71) | -0.30 (1.51) | -0.45 (0.69) |
| Diagnostic Delay | -0.25 (0.27) | -0.13 (0.23) | -0.12 (0.10) |
| Managerial Capacity | 1.74 (1.03) | 1.81 (0.89)* | 0.07 (0.41) |
| Exposures to Job Hazards | -2.72 (0.95)** | -2.54 (0.81)** | -0.18 (0.38) |
| Exposures to Conflict | 0.57 (0.88) | -0.08 (0.77) | 0.66 (0.35) |
| Precision Skills | -0.59 (0.81) | -0.61 (0.71) | -0.09 (0.32) |
| Healthcare | -0.55 (0.75) | -0.32 (0.65) | -0.23 (0.30) |
| Physical Ability | -0.32 (0.88) | -0.37 (0.77) | 0.05 (0.35) |
| Autonomy | 0.36 (0.91) | 0.34 (0.79) | 0.02 (0.36) |
| **B. Worker Characteristics** | **ECAS Total Score** | **ECAS ALS Specific Score** | **ECAS ALS Non-Specific Score** |
| Model R<sup>2</sup> | 0.33 | 0.28 | 0.09 |
| F-statistic | 8.38 | 7.05 | 2.62 |
| DF | 9, 120 | 9, 120 | 9, 120 |
| *P* | 9.19e-10 | 3.05e-8 | .009 |
|  | β (S.E.) | β (S.E.) | β (S.E.) |
| Intercept | 115.17 (7.18)*** | 86.71 (6.25)*** | 28.46 (2.86)*** |
| Age at Test | -0.34 (0.07)*** | -0.27 (0.06)*** | -0.07 (0.03)* |
| Education | 1.04 (0.35)** | 0.76 (0.31)* | 0.29 (0.14)* |
| Non-Bulbar Onset | -0.62 (1.71) | -0.28 (1.49) | -0.34 (0.68) |
| Diagnostic Delay | -0.26 (0.97) | -0.11 (0.23) | -0.14 (0.10) |
| Reasoning Ability | 2.10 (0.97)* | 2.02 (0.84)* | -0.08 (0.39) |
| Visual Perceptual Ability | -1.55 (0.89) | -1.62 (0.77)* | -0.06 (0.30) |
| Social Ability | 1.81 (0.76)* | 1.18 (0.66) | 0.61 (0.30) |
| Coordination Skills | -1.42 (0.76) | -1.05 (0.66) | -0.37 (0.30) |
| Creativity | 1.03 (0.93) | 0.96 (0.81) | 0.08 (0.37) |
| **C. Worker Requirements** | **ECAS Total Score** | **ECAS ALS Specific Score** | **ECAS ALS Non-Specific Score** |
| Model R<sup>2</sup> | 0.34 | 0.30 | 0.09 |
| F-statistic | 8.90 | 7.67 | 2.48 |
| DF | 9, 120 | 9, 120 | 9, 120 |
| *p* | 2.44e-10 | 5.90e-10 | .012 |
|  | β (S.E.) | β (S.E.) | β (S.E.) |
| Intercept | 117.68 (7.18)*** | 88758 (6.23)*** | 28.41 (2.88)*** |
| Age at Test | -0.35 (0.07)*** | -0.28 (0.06)*** | -0.07 (0.03)* |
| Education | 0.94 (0.35)** | 0.68 (0.30)* | 0.26 (0.14) |
| Non-Bulbar Onset | -0.36 (1.71) | -0.08 (1.48) | -0.27 (0.69) |
| Diagnostic Delay | -0.32 (0.26) | -0.18 (0.23) | -0.14 (0.10) |
| Analytical Skills | 3.25 (1.12)** | 3.14 (0.98)** | -0.11 (0.45) |
| Technical Skills | -2.29 (0.78)** | -1.84 (0.67)** | 0.44 (0.31) |
| Health Services Knowledge | 0.91 (0.76) | 0.57 (0.66) | -0.34 (0.31) |
| Business skills/Knowledge | 0.42 (0.62) | 0.15 (0.54) | 0.26 (0.25) |
| Humanities Knowledge | 1.93 (0.70)** | 1.51 (0.61)* | -0.43 (0.28) |

\* p < .05; \*\* p < .01; \*\*\* p < .001; ECAS = Edinburgh Cognitive and Behavioral ALS Screen

**Table 3.** Multiple linear regression models of motor dysfunction with O*NET Occupation Requirements (A), Worker Characteristics (B), and Worker Requirements (C)

| **A. Occupational Requirements** | **UMN Total Score** | **ALSFRS-R Total Score** |
| --- | --- | --- |
| Model R<sup>2</sup> | 0.15 | 0.04 |
| F-statistic | 3.05 | 1.43 |
| DF | 11, 126 | 11, 119 |
| *p* | .003 | .026 |
| | $\beta$ (S.E.) | $\beta$ (S.E.) |
| Intercept | 30.28 (7.15)*** | 35.58 (5.67)*** |
| Age at Test | -0.19 (0.08)* | -0.03 (0.06) |
| Education | -0.72 (0.35)* | 0.26 (0.27) |
| Non-Bulbar Onset | 0.97 (1.78) | -3.61 (1.46)** |
| Diagnostic Delay | -0.17 (0.34) | -0.34 (0.24) |
| Managerial Capacity | 1.06 (1.02) | -0.88 (0.84) |
| Exposures to job hazards | -1.63 (0.93) | 0.35 (0.79) |
| Exposures to conflict | 0.52 (0.87) | -1.15 (0.77) |
| Precision skills | 1.86 (0.81)* | 0.02 (0.71) |
| Healthcare | 1.27 (0.74) | -0.77 (0.61) |
| Physical ability | 0.45 (0.80) | 0.28 (0.73) |
| Autonomy | 1.11 (0.96) | -0.62 (0.77) |
| **B. Worker Characteristics** | **UMN Total Score** | **ALSFRS-R Total Score** |
| Model R<sup>2</sup> | 0.08 | 0.04 |
| F-statistic | 2.07 | 1.62 |
| DF | 9, 128 | 9, 128 |
| *p* | .038 | .011 |
| | $\beta$ (S.E.) | $\beta$ (S.E.) |
| Intercept | 30.96 (7.30)*** | 36.64 (5.61)*** |
| Age at Test | -0.22 (0.08)** | -0.03 (0.06) |
| Education | -0.54 (0.36) | 0.23 (0.27) |
| Non-Bulbar Onset | 0.86 (1.85) | -3.70 (1.45)** |
| Diagnostic Delay | -0.15 (0.36) | -0.28 (0.23) |
| Reasoning Ability | 2.13 (0.98)* | -0.58 (0.74) |
| Visual-perceptual Ability | -1.50 (0.87) | 0.27 (0.73) |
| Social ability | 0.75 (0.80) | -0.99 (0.62) |
| Coordination skills | 1.40 (0.76) | -0.19 (0.61) |
| Creativity | 0.11 (0.99) | -0.66 (0.79) |
| **C. Worker Requirements** | **UMN Total Score** | **ALSFRS-R Total Score** |
| Model R<sup>2</sup> | 0.04 | 0.07 |
| F-statistic | 1.67 | 2.243 |
| DF | 9, 128 | 9, 128 |
| *p* | .016 | .033 |
| | $\beta$ (S.E.) | $\beta$ (S.E.) |
| Intercept | 26.02 (7.69)*** | 37.32 (5.66)*** |
| Age at Test | -0.21 (0.08)** | -0.03 (0.06) |
| Education | -0.45 (0.37) | 0.18 (0.26) |
| Non-Bulbar Onset | 1.08 (1.92) | -4.09(1.46)** |
| Diagnostic Delay | -0.26 (0.38) | -0.25 (0.24) |
| Analytical skills | 2.23 (1.18) | -0.11 (0.89) |
| Technical skills | -0.93 (0.85) | 0.54 (0.63) |
| Health services knowledge | 1.10 (0.86) | -1.41 (0.61)* |
| Business skill | 0.22 (0.64) | -0.52 (0.48) |
| Humanities knowledge | 0.47 (0.74) | -1.01 (0.55)* |

\* p < .05; \*\* p < .01; \*\*\* p < .001; PUMNS = Penn Upper Motor Neuron Scale; ALSFRS-R = ALS Functional Rating Scale - Revised

**Figure 1.**
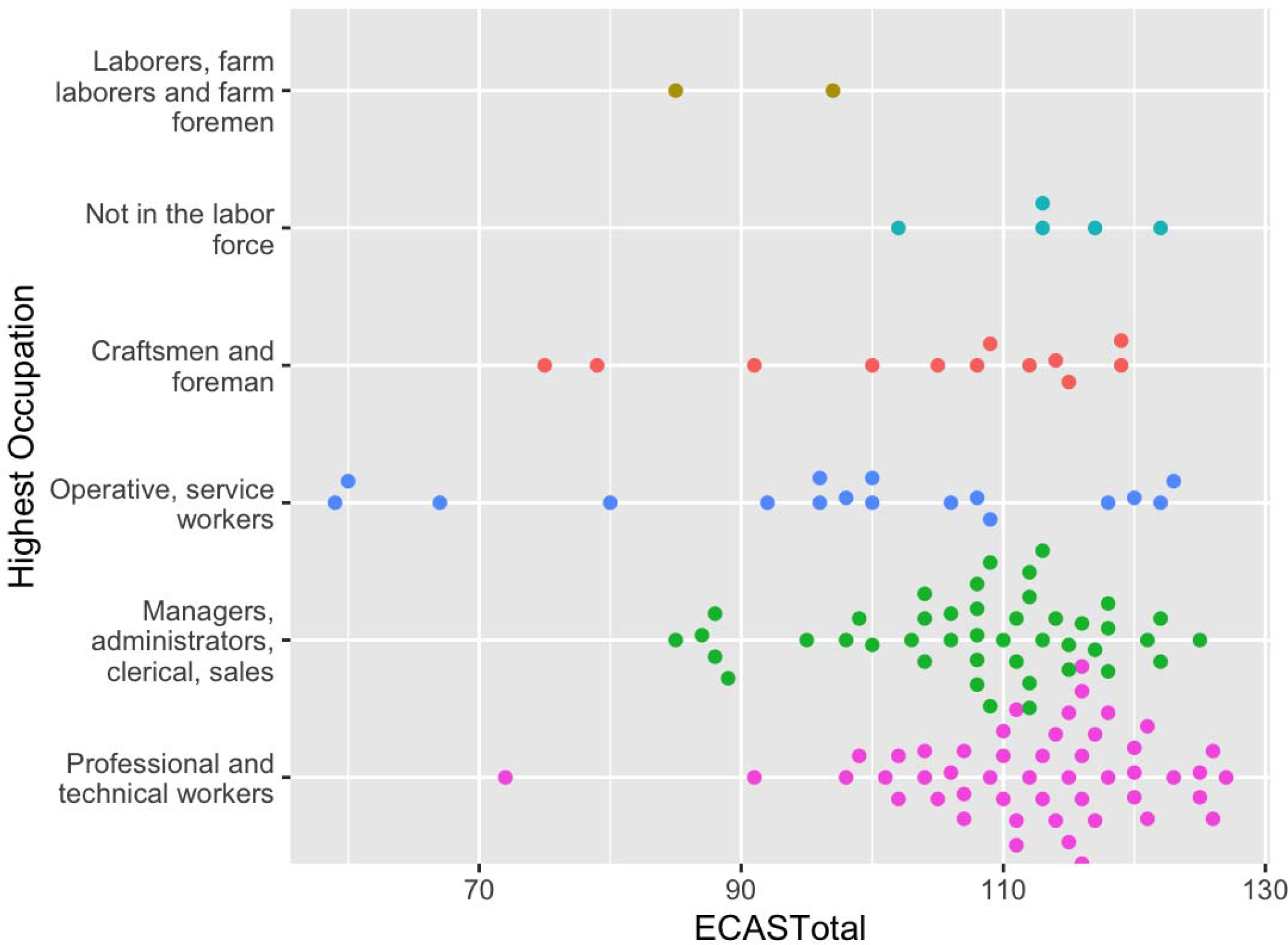
Beeswarm plot of ECAS Total Scores by primary occupation categories

### Impact of Occupation Features on Cognitive Performance

#### Occupational Requirements

After covarying for age, education, diagnostic delay, and ALS symptom onset site (bulbar vs. non-bulbar), a history of working in occupations with greater likelihood of Exposures to Job Hazards was associated with poorer performance on the ECAS Total (β = -2.72, *p* = 0.007) and ECAS ALS Specific scores (β = -2.54, *p* = 0.002). Occupations requiring Managerial Capacity were associated with better performance on the ECAS ALS-Specific score (β = 1.81, *p* = 0.043). No occupational requirement factors were associated with ECAS ALS Non-Specific scores (all *p*-values > .05).

#### Worker Characteristics

After covarying for age, education, diagnostic delay, and ALS symptom onset site, a history of working in occupations that demand greater Reasoning Ability was associated with better performance on the ECAS Total (β = 2.10, *p* = 0.029) and ECAS ALS Specific scores (β = 2.02, *p* = 0.016). Occupations that involve greater Social Ability were associated with better performance on the ECAS Total score (β = 1.81, *p* = 0.024). Occupations that rely on Visual Perceptual Ability were associated with worse performance on the ECAS ALS Specific score (β = -1.62, *p* = 0.041). No worker characteristic factors were associated with ECAS ALS Non-Specific scores (all *p*-values > .05) and the omnibus model did not survive correction for multiple comparisons (p > .005).

#### Worker Requirements

After covarying for age, education, diagnostic delay, and ALS symptom onset site, a history of working occupations that require more Analytical Skills was associated with better performance on the ECAS Total (β = 3.25, *p* = 0.006) and ALS Specific scores (β = 3.14, *p* = 0.002). Working occupations that require more Humanities Knowledge was associated with better performance on the ECAS Total (β = 1.93, *p* = 0.009) and ALS Specific scores (β = 1.51, *p* = 0.017). Working jobs that require more Technical Skills was associated with worse performance on the ECAS Total (β = -2.29, *p* = 0.006) and ALS Specific scores (β = -1.84, *p* = 0.009). No worker requirement factors were associated with ECAS ALS Non-Specific scores (all *p*-values > .05) and the omnibus model did not survive correction for multiple comparisons (p > .005).

### Impact of Occupation Features on Motor Impairment

#### Occupational Requirements

After covarying for age, education, diagnostic delay, and ALS symptom onset site, a history of working an occupation with greater likelihood of requiring Precision Skills was associated with greater motor impairment on the PUMNS (β = 1.86, *p* = .018). No occupational requirements were associated with FRS Total Scores (all *p*-values > .05), and the omnibus model was not significant following correction for multiple comparisons (*p* > .005).

#### Worker Characteristics

After covarying for age, education, diagnostic delay, and ALS symptom onset site, omnibus models for the PUMNS and ALSFRS-R were not significant following correction for multiple comparisons (*p*-values > .005).

#### Worker Requirements

After covarying for age, education, diagnostic delay, and ALS symptom onset site, omnibus models for the PUMNS and ALSFRS-R were not significant following correction for multiple comparisons (*p*-values > .005).

## Discussion

This study was conducted to assess the impact of empirically derived occupational factors on cognitive and motor functioning in ALS. Our results show that specific occupational skills and requirements can have a protective effect on cognitive functioning while others can confer elevated risk for cognitive dysfunction. We found that skills and requirements typically found in white collar jobs, such as managerial capacity, reasoning ability, and analytic skills were linked to better performance on the ECAS, while job factors representing hazardous working conditions and less cognitively demanding activities (i.e., technical skills, visual-perceptual ability) were associated with worse performance on the ECAS. Associations between occupational factors and motor dysfunction were less robust but showed evidence for an increased risk for motor dysfunction related to jobs that require repetitive and precise tasks. Each of these findings will be discussed below.

The first aim of our study was to assess the impact of occupational skills and requirements on cognitive functioning in ALS. We found a protective effect of managerial capacity, reasoning ability, social ability, analytical skills, and humanities knowledge. These skills and abilities are featured in more complex jobs that are typically more cognitively demanding and require higher educational attainment. For example, O*NET job categories highest in analytical skills (i.e., those that involve complex problem solving and the ability to learn new skills) include scientists, engineers, and attorneys, while jobs lowest in analytic skills include construction laborers, street vendors, and parking attendants. This finding is consistent with the extant literature showing a protective effect of occupational complexity on cognition in healthy aging,^17^ dementia,^14, 31^ and ALS^8, 13^ and highlights the importance of mid-life contributions to CR, which are thought to maintain cognitive abilities via enhanced neural activity secondary to continued use of higher order cognitive skills. However, unlike other studies supporting the “use it or lose it hypothesis,” education was also a significant predictor of better cognitive performance even after accounting for occupational skills and requirements, suggesting that CR likely reflects a combination of static and dynamic features of an individual’s personal history. Additionally, we found a deleterious effect of exposure to job hazards, visual perceptual ability, and technical skills on cognitive functioning in ALS. Exposures to job hazards includes varying circumstances that put employees at risk (e.g., extreme temperatures, environmental neurotoxicants and mutagens, operating heavy machinery) and has been identified as a risk factor for developing sporadic ALS^32–35^ and cognitive decline in other populations.^36–38^ Exposure to job hazards disproportionately affects minoritized individuals who are more likely to encounter unsafe working conditions,^38^ and may account for reports of lower cognitive functioning in African American ALS patients even after application of demographic normative correction.^39^ Further, visuo-perceptual ability (i.e., spatial orientation) and technical skills (i.e., mechanical knowledge) may be more essential in blue collar jobs that do not rely on high order cognitive skills. For example, worker characteristics with high factor loadings for Visuo-perceptual ability included peripheral vision (0.88) and spatial orientation abilities (0.87), which occur at the highest levels among O*NET job categories of bus and truck drivers and other machine operators (cranes, tractors, trains) and lowest among social service and healthcare workers (probation officers, occupational therapists, medical assistants; see Supplemental Tables 1-3). Our study improves upon previous findings by providing more a nuanced assessment of occupational factors as they relate to cognitive reserve in ALS.

Our second aim was to assess the impact of specific occupational skills and requirements on motor functioning in ALS. Consistent with prior literature demonstrating increased risk for ALS among individuals with a history of greater physical activity,^18–20^ we found that occupational requirements showed an increased risk for motor impairment. Specifically, we observed that a history of working jobs that require precision skills, characterized by repeatedly performing the same task with high precision (e.g., welder or assembly line worker), was linked to worse motor functioning on the PUMNS. This may reflect increased vulnerability to motor impairment in ALS due to overuse of motor skills among individuals who worked jobs with high motor demands. The direction of this finding runs contrary to a previous study that reported attenuated motor dysfunction among individuals with higher CR but only for those with bulbar onset.^8^ In contrast, our results did not show evidence for the motor reserve hypothesis in ALS despite using the same outcome measure (ALSFRS-R), which may reflect a stronger effect of non-occupational CR proxies that were included in the previous study but not reflected in our analyses (bilingualism and midlife leisure activities). Notably, only one model for this aim survived correction for multiple comparisons and all models accounted for a much smaller proportion of the overall variance in motor functioning relative to cognitive performance, suggesting that occupational history has a greater influence on cognitive relative to motor functioning in ALS.

The strengths of our study include the use of a large, well-characterized sample of ALS patients and use of O*NET categorization to provide a more detailed assessment of occupational complexity. Nonetheless, our findings are limited by several factors. First, we did not have a healthy control group to compare against, so it remains unclear if our findings are specifc to ALS or represent a more general protective effect of CR on cognitive functioning across the lifespan. Studies of CR assessing occupational complexity in healthy aging have shown protective effects of occupational cognitive complexity,^40^ mental demands at work,^41^ and enriched work environments^42^ defined using O*NET-derived measures on level and slope of global cognition and episodic memory over time. In contrast, we did not find an effect of any O*NET factors on the ECAS ALS Non-Specific Score, which includes measurement of episodic memory. Instead, all of our significant CR findings were reflected in the ECAS ALS-Specific Score, suggesting that our results may reflect preservation of cognitive abilities most vulnerable to impairment in ALS (i.e., language, executive functions, verbal fluency), as opposed to a episodic memory, which is most likely to be affected in health aging samples. Fewer studies have investigated the impact of occupation on language and executive functioning performance, but limited evidence suggests that specific occupations (e.g., teacher) and job characteristics (e.g., demand, control) can contribute to better verbal fluency performance in healthy samples.^43, 44^ Taken together, it remains unclear if our findings reflect a specific effect of occupation on cognitive performance in ALS, and future research should include case controlled studies to address this gap. Additional limitations include the use of cross-sectional data, which limits our ability to draw longitudinal conclusions about cognitive trajectories in ALS, and the demographics of our sample, which was highly educated with a predominantly white sample lacking diversity. Similarly, the distribution of the occupations in our sample is disproportionately represented by white collar workers (i.e. managers, administrators), which limits the generalizability of our findings.

This study provides additional support for the CR hypothesis in ALS and shows that specific occupational skills and requirements can have a protective effect on cognitive functioning in ALS, while others may impart increased risk for cognitive impairment. In contrast, we did not find evidence to support a motor reserve hypothesis in ALS, as all associations between motor symptoms and occupational factors showed an increased risk for motor dysfunction. Taken together, our findings highlight the role of occupation as a salient CR proxy that should be considered a significant contributor to clinical outcomes in ALS.

## Data Availability

All data produced in the present study are available upon reasonable request to the authors

## References

1. Phukan J, Elamin M, Bede P, et al. The syndrome of cognitive impairment in amyotrophic lateral sclerosis: a population-based study. J Neurol Neurosurg Psychiatry. 2012;83(1):102–108. doi:10.1136/jnnp-2011-300188

2. McMillan CT. Defining cognitive impairment in amyotrophic lateral sclerosis: an evaluation of empirical approaches. Published 2022. Accessed April 4, 2023. https://www.tandfonline.com/doi/epub/10.1080/21678421.2022.2039713?needAccess=true

3. Yang T. Risk factors for cognitive impairment in amyotrophic lateral sclerosis: a systematic review and meta-analysis | Journal of Neurology, Neurosurgery & Psychiatry. Published 2021. Accessed April 4, 2023. https://jnnp.bmj.com/content/92/7/688.abstract

4. Canosa A, Palumbo F, Iazzolino B, et al. The interplay among education, brain metabolism, and cognitive impairment suggests a role of cognitive reserve in Amyotrophic Lateral Sclerosis. Neurobiology of Aging. 2021;98:205–213. doi:10.1016/j.neurobiolaging.2020.11.010

5. Temp AGM, Prudlo J, Vielhaber S, et al. Cognitive reserve and regional brain volume in amyotrophic lateral sclerosis. Cortex. 2021;139:240–248. doi:10.1016/j.cortex.2021.03.005

6. Temp AGM, Kasper E, Machts J, et al. Cognitive reserve protects ALS-typical cognitive domains: A longitudinal study. Annals of Clinical and Translational Neurology. 2022;9(8):1212–1223. doi:10.1002/acn3.51623

7. Placek K, Massimo L, Olm C, et al. Cognitive reserve in frontotemporal degeneration: Neuroanatomic and neuropsychological evidence. Neurology. 2016;87(17):1813–1819. doi:10.1212/WNL.0000000000003250

8. Consonni M, Dalla Bella E, Bersano E, Telesca A, Lauria G. Cognitive reserve is associated with altered clinical expression in amyotrophic lateral sclerosis. Amyotrophic Lateral Sclerosis and Frontotemporal Degeneration. 2021;22(3-4):237–247. doi:10.1080/21678421.2020.1849306

9. Stern Y. Cognitive reserveL. Neuropsychologia. 2009;47(10):2015-2028. doi:10.1016/j.neuropsychologia.2009.03.004

10. Stern Y. Cognitive reserve in ageing and Alzheimer’s disease. The Lancet Neurology. 2012;11(11):1006–1012. doi:10.1016/S1474-4422(12)70191-6

11. Guzzetti S, Mancini F, Caporali A, Manfredi L, Daini R. The association of cognitive reserve with motor and cognitive functions for different stages of Parkinson’s disease. Experimental Gerontology. 2019;115:79–87. doi:10.1016/j.exger.2018.11.020

12. Stern Y, Arenaza-Urquijo EM, Bartrés-Faz D, et al. Whitepaper: Defining and investigating cognitive reserve, brain reserve, and brain maintenance. Alzheimer’s & Dementia. 2020;16(9):1305–1311. doi:10.1016/j.jalz.2018.07.219

13. Costello E, Rooney J, Pinto-Grau M, et al. Cognitive reserve in amyotrophic lateral sclerosis (ALS): a population-based longitudinal study. J Neurol Neurosurg Psychiatry. 2021;92(5):460–465. doi:10.1136/jnnp-2020-324992

14. Placek K, Massimo L, Olm C, et al. Cognitive reserve in frontotemporal degeneration. Neurology. 2016;87(17):1813–1819. doi:10.1212/WNL.0000000000003250

15. Suo C, León I, Brodaty H, et al. Supervisory experience at work is linked to low rate of hippocampal atrophy in late life. NeuroImage. 2012;63(3):1542–1551. doi:10.1016/j.neuroimage.2012.08.015

16. Dekhtyar S, Wang HX, Scott K, Goodman A, Koupil I, Herlitz A. A Life-Course Study of Cognitive Reserve in Dementia—From Childhood to Old Age. The American Journal of Geriatric Psychiatry. 2015;23(9):885–896. doi:10.1016/j.jagp.2015.02.002

17. Andel R, Kåreholt I, Parker MG, Thorslund M, Gatz M. Complexity of Primary Lifetime Occupation and Cognition in Advanced Old Age. J Aging Health. 2007;19(3):397–415. doi:10.1177/0898264307300171

18. Daneshvar DH, Mez J, Alosco ML, et al. Incidence of and Mortality From Amyotrophic Lateral Sclerosis in National Football League Athletes. JAMA Netw Open. 2021;4(12):e2138801. doi:10.1001/jamanetworkopen.2021.38801

19. Julian TH, Glascow N, Barry ADF, et al. Physical exercise is a risk factor for amyotrophic lateral sclerosis: Convergent evidence from Mendelian randomisation, transcriptomics and risk genotypes. EBioMedicine. 2021;68:103397. doi:10.1016/j.ebiom.2021.103397

20. Chapman L, Cooper-Knock J, Shaw PJ. Physical activity as an exogenous risk factor for amyotrophic lateral sclerosis: a review of the evidence. Brain. 2023;146(5):1745–1757. doi:10.1093/brain/awac470

21. Brooks BR, Miller RG, Swash M, Munsat TL. El Escorial revisited: Revised criteria for the diagnosis of amyotrophic lateral sclerosis. Amyotrophic Lateral Sclerosis and Other Motor Neuron Disorders. 2000;1(5):293–299. doi:10.1080/146608200300079536

22. Strong MJ, Abrahams S, Goldstein LH, et al. Amyotrophic lateral sclerosis - frontotemporal spectrum disorder (ALS-FTSD): Revised diagnostic criteria. Amyotrophic Lateral Sclerosis and Frontotemporal Degeneration. 2017;18(3-4):153–174. doi:10.1080/21678421.2016.1267768

23. Abrahams S, Newton J, Niven E, Foley J, Bak TH. Screening for cognition and behaviour changes in ALS. Amyotrophic Lateral Sclerosis and Frontotemporal Degeneration. 2014;15(1-2):9–14. doi:10.3109/21678421.2013.805784

24. Cedarbaum JM, Stambler N, Malta E, et al. The ALSFRS-R: a revised ALS functional rating scale that incorporates assessments of respiratory function. Journal of the Neurological Sciences. 1999;169(1-2):13–21. doi:10.1016/S0022-510X(99)00210-5

25. Quinn C, Edmundson C, Dahodwala N, Elman L. Reliable and efficient scale to assess upper motor neuron disease burden in amyotrophic lateral sclerosis. Muscle Nerve. 2020;61(4):508–511. doi:10.1002/mus.26764

26. Cifuentes M, Boyer J, Lombardi DA, Punnett L. Use of O*NET as a job exposure matrix: A literature review. Am J Ind Med. Published online 2010:n/a-n/a. doi:10.1002/ajim.20846

27. Gadermann AM, Heeringa SG, Stein MB, et al. Classifying U.S. Army Military Occupational Specialties Using the Occupational Information Network. Military Medicine. 2014;179(7):752–761. doi:10.7205/MILMED-D-13-00446

28. Hadden WC, Kravets N, Muntaner C. Descriptive dimensions of US occupations with data from the O*NET. Social Science Research. 2004;33(1):64–78. doi:10.1016/S0049-089X(03)00039-5

29. Peterson NG, Mumford MD, Borman WC, Jeanneret PR, Fleishman EA. An Occupational Information System for the 21st Century: The Development of O*NET. American Psychological Association; 1999:xii, 336. doi:10.1037/10313-000

30. Spreng RN, Rosen HJ, Strother S, et al. Occupation attributes relate to location of atrophy in frontotemporal lobar degeneration. Neuropsychologia. 2010;48(12):3634–3641. doi:10.1016/j.neuropsychologia.2010.08.020

31. Smyth KA, Fritsch T, Cook TB, McClendon MJ, Santillan CE, Friedland RP. Worker functions and traits associated with occupations and the development of AD. Neurology. 2004;63(3):498–503. doi:10.1212/01.WNL.0000133007.87028.09

32. Dickerson AS, Hansen J, Specht AJ, Gredal O, Weisskopf MG. Population-based study of amyotrophic lateral sclerosis and occupational lead exposure in Denmark. Occup Environ Med. 2019;76(4):208–214. doi:10.1136/oemed-2018-105469

33. Dickerson AS, Hansen J, Gredal O, Weisskopf MG. Amyotrophic Lateral Sclerosis and Exposure to Diesel Exhaust in a Danish Cohort. American Journal of Epidemiology. 2018;187(8):1613–1622. doi:10.1093/aje/kwy069

34. Dickerson AS, Hansen J, Kioumourtzoglou MA, Specht AJ, Gredal O, Weisskopf MG. Study of occupation and amyotrophic lateral sclerosis in a Danish cohort. Occup Environ Med. 2018;75(9):630–638. doi:10.1136/oemed-2018-105110

35. Visser AE, D’Ovidio F, Peters S, et al. Multicentre, population-based, case–control study of particulates, combustion products and amyotrophic lateral sclerosis risk. J Neurol Neurosurg Psychiatry. 2019;90(8):854–860. doi:10.1136/jnnp-2018-319779

36. Crous-Bou M, Gascon M, Gispert JD, et al. Impact of urban environmental exposures on cognitive performance and brain structure of healthy individuals at risk for Alzheimer’s dementia. Environment International. 2020;138:105546. doi:10.1016/j.envint.2020.105546

37. Cabral Pinto MMS, Marinho-Reis AP, Almeida A, et al. Human predisposition to cognitive impairment and its relation with environmental exposure to potentially toxic elements. Environ Geochem Health. 2018;40(5):1767–1784. doi:10.1007/s10653-017-9928-3

38. Friedman-Jimenez G. Occupational disease among minority workers: a common and preventable public health problem. AAOHN Journal. Published online 1989.

39. Yancey J. The Prevalence and One Year Incidence of Frontotemporal Dementia in Individuals with Amyotrophic Lateral Sclerosis Based on Race. University of Pennsylvania; 2020. https://www.proquest.com/docview/2418740295?pq-origsite=gscholar&fromopenview=true

40. Pool LR, Weuve J, Wilson RS, Bültmann U, Evans DA, Mendes de Leon CF. Occupational cognitive requirements and late-life cognitive aging. Neurology. 2016;86(15):1386–1392. doi:10.1212/WNL.0000000000002569

41. Fisher GG, Stachowski A, Infurna FJ, Faul JD, Grosch J, Tetrick LE. Mental work demands, retirement, and longitudinal trajectories of cognitive functioning. Journal of Occupational Health Psychology. 2014;19(2):231–242. doi:10.1037/a0035724

42. Then FS, Luck T, Luppa M, Konig HH, Angermeyer MC, Riedel-Heller SG. Differential effects of enriched environment at work on cognitive decline in old age. Neurology. 2015;84(21):2169–2176. doi:10.1212/WNL.0000000000001605

43. Sabbath EL, Andel R, Zins M, Goldberg M, Berr C. Domains of cognitive function in early old age: which ones are predicted by pre-retirement psychosocial work characteristics? Occup Environ Med. 2016;73(10):640–647. doi:10.1136/oemed-2015-103352

44. Van der Elst W, Van Boxtel MPJ, Jolles J. Occupational Activity and Cognitive Aging: A Case-Control Study Based on the Maastricht Aging Study. Experimental Aging Research. 2012;38(3):315–329. doi:10.1080/0361073X.2012.672137

