## Supplemental Tables for "Cognitive reserve in ALS: The role of occupational skills and requirements"

**Supplemental Table 1.** Factor Loadings with Varimax Rotation for O*NET Occupational Requirements

|  | Managerial Capacity | Exposure to Job Hazards | Exposure to Conflict | Precision Skills | Healthcare | Physical Ability | Autonomy |
| --- | --- | --- | --- | --- | --- | --- | --- |
| Operating Vehicles Mechanized Devices or Equipment | 0.043505 | 0.897906 | 0.012768 | -0.01778 | 0.017383 | 0.006371 | -0.02 |
| Very Hot or Cold Temperatures | 0.159124 | 0.892399 | 0.011801 | 0.034169 | -0.16768 | 0.006205 | -0.10308 |
| Extremely Bright or Inadequate Lighting | 0.135894 | 0.86869 | 0.055058 | -0.01884 | 0.010617 | 0.055637 | -0.05571 |
| Exposed to Hazardous Equipment | 0.137966 | 0.860467 | -0.19687 | -0.1558 | 0.055754 | -0.13909 | -0.06223 |
| Outdoors Exposed to Weather | 0.014057 | 0.839732 | 0.14714 | 0.163298 | -0.2138 | 0.234423 | 0.089954 |
| Indoors Not Environmentally Controlled | 0.008613 | 0.837 | -0.11299 | -0.01302 | -0.18033 | 0.040157 | -0.02386 |
| Exposed to High Places | 0.036087 | 0.823654 | -0.04734 | -0.04857 | -0.0908 | 0.050212 | -0.00628 |
| In an Open Vehicle or Equipment | 0.080932 | 0.81897 | -0.07879 | -0.0123 | -0.15386 | 0.050988 | -0.0634 |
| Performing General Physical Activities | 0.159122 | 0.811109 | 0.100972 | 0.140248 | 0.140986 | -0.31966 | -0.02756 |
| Cramped Work Space Awkward Positions | 0.167464 | 0.810658 | 0.049529 | -0.01628 | 0.224599 | -0.13032 | 0.007682 |
| Spend Time Climbing Ladders Scaffolds or Poles | 0.059333 | 0.783254 | -0.08276 | -6.53E-05 | -0.11337 | -0.03171 | 0.023367 |
| Exposed to Minor Burns Cuts Bites or Stings | 0.26594 | 0.781194 | -0.06922 | 0.015662 | 0.095163 | -0.28554 | -0.02521 |
| Repairing and Maintaining Mechanical Equipment | 0.084166 | 0.779814 | -0.2793 | -0.14016 | 0.214577 | -0.19481 | 0.017137 |
| Outdoors Under Cover | -0.06522 | 0.776062 | 0.078428 | 0.149171 | -0.18289 | 0.232991 | 0.127895 |
| Exposed to Contaminants | 0.278016 | 0.770004 | -0.03827 | -0.09935 | 0.236605 | -0.21914 | -0.11261 |
| Exposed to Whole Body Vibration | 0.089138 | 0.763034 | -0.03605 | -0.00901 | -0.0745 | 0.042013 | -0.09094 |
| Exposed to Hazardous Conditions | 0.050538 | 0.75283 | -0.16149 | -0.16468 | 0.31483 | -0.08932 | -0.04634 |
| Wear Common Protective or Safety | 0.113889 | 0.750944 | -0.12309 | -0.14255 | 0.320036 | -0.29717 | -0.12095 |
| Sounds Noise Levels Are Distracting or Uncomfortable | 0.203387 | 0.72549 | 0.025381 | -0.24864 | 0.037922 | -0.09304 | -0.26695 |
| Spend Time Keeping or Regaining Balance | 0.209619 | 0.716766 | 0.129607 | 0.15655 | 0.020594 | -0.29666 | -0.06606 |
| Handling and Moving Objects | 0.276004 | 0.700031 | -0.03657 | 0.012404 | 0.227427 | -0.43293 | -0.04544 |
| In an Enclosed Vehicle or Equipment | -0.08311 | 0.689353 | 0.197719 | 0.119278 | -0.22004 | 0.372315 | 0.23555 |
| Wear Specialized Protective or Safety Equipment such as Breathing Apparatus Safety Harness Full Protection Suits or Radiation Protection | -0.02853 | 0.687968 | -0.01481 | -0.06937 | 0.342859 | -0.17855 | -0.04131 |
| Inspecting Equipment Structures or Material | -0.15462 | 0.686929 | -0.11152 | -0.16126 | 0.432289 | -0.15208 | -0.07588 |
| Controlling Machines and Processes | 0.108789 | 0.683335 | -0.2607 | -0.22868 | 0.36218 | -0.29259 | -0.08443 |
| Spend Time Kneeling Crouching Stooping or Crawling | 0.270849 | 0.678616 | 0.041622 | 0.148094 | 0.071749 | -0.33819 | 0.015582 |
| Responsible for Others Health and Safety | -0.09383 | 0.66458 | 0.32158 | -0.17737 | 0.183433 | -0.3435 | -0.13585 |
| Spend Time Bending or Twisting the Body | 0.417225 | 0.627237 | 0.079148 | 0.061899 | 0.147265 | -0.49426 | -0.1067 |
| Spend Time Walking and Running | 0.323017 | 0.530258 | 0.209993 | 0.071609 | 0.044994 | -0.50695 | -0.22793 |
| Pace Determined by Speed of Equipment | 0.308432 | 0.513065 | -0.21931 | -0.32542 | 0.053607 | -0.26694 | -0.39334 |
| Repairing and Maintaining Electronic Equipment | -0.0739 | 0.510939 | -0.33366 | -0.19793 | 0.331382 | -0.05726 | 0.045002 |
| Work Schedules | 0.039211 | 0.486031 | -0.10322 | 0.309201 | -0.20792 | 0.002918 | 0.217025 |
| Spend Time Standing | 0.352853 | 0.465381 | 0.095897 | 0.147375 | 0.091526 | -0.64214 | -0.13211 |
| Spend Time Using Your Hands to Handle Control or Feel Objects Tools or Controls | 0.516162 | 0.461511 | -0.1635 | -0.1566 | 0.28533 | -0.37266 | -0.01419 |
| Drafting Laying Out and Specifying Technical Devices Parts and Equipment | -0.32366 | 0.46015 | -0.40795 | -0.16141 | 0.09919 | -0.05715 | 0.065708 |
| Consequence of Error | -0.10202 | 0.450891 | 0.259126 | -0.34669 | 0.471842 | 0.054522 | -0.00748 |
| Responsibility for Outcomes and Results | -0.42963 | 0.328753 | 0.211808 | -0.49408 | -0.0935 | -0.31342 | 0.070912 |
| Monitor Processes Materials or Surroundings | -0.56923 | 0.301216 | 0.105261 | -0.11152 | 0.576136 | 0.037628 | -0.08032 |
| Duration of Typical Work Week | -0.55275 | 0.254569 | -0.1962 | -0.30167 | -0.13572 | 0.209718 | 0.036954 |
| Estimating the Quantifiable Characteristics of Products Events or Information | -0.71099 | 0.194408 | -0.27536 | -0.0877 | 0.219617 | 0.085049 | 0.007168 |
| Impact of Decisions on Co workers or Company Results | -0.28523 | 0.17074 | 0.478235 | -0.38183 | 0.133094 | 0.120394 | 0.400347 |
| Frequency of Decision Making | -0.06107 | 0.169245 | 0.585522 | -0.37696 | 0.114634 | 0.045276 | 0.375587 |
| Spend Time Making Repetitive Motions | 0.560459 | 0.138975 | -0.08631 | -0.2528 | 0.053395 | -0.2876 | -0.18793 |
| Exposed to Radiation | -0.07484 | 0.122861 | 0.179842 | -0.08573 | 0.585461 | -0.10397 | 0.023991 |
| Coordinating the Work and Activities of Others | -0.87142 | 0.085618 | 0.129475 | -0.05092 | -0.15789 | -0.1485 | -0.00331 |
| Time Pressure | -0.03226 | 0.080307 | 0.070746 | -0.5901 | -0.04629 | -0.03384 | 0.103735 |
| Deal With Physically Aggressive People | -0.03712 | 0.080121 | 0.737356 | 0.140797 | 0.169839 | 0.043252 | -0.21286 |
| Physical Proximity | 0.147427 | 0.074189 | 0.592593 | 0.071666 | 0.347649 | -0.39518 | -0.02873 |
| Monitoring and Controlling Resources | -0.73427 | 0.04994 | -0.00388 | -0.09302 | -0.06462 | -0.12917 | 0.273411 |
| Guiding Directing and Motivating Subordinates | -0.87331 | 0.045586 | 0.143138 | -0.05542 | -0.11864 | -0.1993 | 0.021099 |
| Coordinate or Lead Others | -0.50121 | 0.034922 | 0.447108 | -0.32686 | -0.09827 | -0.1185 | 0.02957 |
| Face to Face Discussions | -0.37757 | 0.032171 | 0.329373 | -0.28982 | 0.044664 | 0.029344 | 0.165007 |
| Evaluating Information to Determine Compliance with Standards | -0.65464 | 0.026849 | 0.162655 | -0.21549 | 0.324856 | 0.278331 | -0.10911 |
| Work With Work Group or Team | -0.38445 | 0.026316 | 0.45995 | -0.40333 | -0.03713 | -0.12299 | -0.21897 |
| Judging the Qualities of Things Services or People | -0.77648 | 0.004008 | 0.027054 | -0.01247 | 0.164906 | 0.022244 | 0.103445 |
| Developing and Building Teams | -0.88205 | 0.00069 | 0.172486 | -0.0659 | -0.0967 | -0.11097 | -0.09229 |
| Assisting and Caring for Others | -0.23524 | -0.01713 | 0.652005 | 0.1653 | 0.439528 | -0.2482 | 0.023546 |
| Frequency of Conflict Situations | -0.24736 | -0.02161 | 0.757021 | -0.22995 | -0.10183 | 0.061887 | -0.00409 |
| Deal With Unpleasant or Angry People | 0.101519 | -0.04984 | 0.823778 | -0.14078 | 0.043446 | -0.00863 | -0.09708 |
| Making Decisions and Solving Problems | -0.8285 | -0.05384 | 0.036334 | -0.05734 | 0.272107 | 0.175199 | 0.134461 |
| Level of Competition | -0.43309 | -0.05627 | -0.12928 | -0.19817 | -0.12391 | -0.07536 | 0.359287 |
| Scheduling Work and Activities | -0.87449 | -0.05825 | 0.015274 | 0.047898 | -0.07345 | 0.01754 | 0.163552 |
| Training and Teaching Others | -0.80202 | -0.06211 | 0.096798 | 0.145768 | 0.12684 | -0.09108 | -0.05784 |
| Freedom to Make Decisions | -0.38399 | -0.06881 | 0.117176 | 0.075299 | 0.089528 | 0.15913 | 0.636578 |
| Degree of Automation | 0.097141 | -0.07023 | -0.04041 | -0.56137 | -0.01992 | 0.237683 | -0.36727 |
| Importance of Repeating Same Tasks | 0.261472 | -0.07313 | 0.189219 | -0.62014 | 0.204664 | 0.168466 | -0.14092 |
| Exposed to Disease or Infections | -0.06072 | -0.09207 | 0.57511 | 0.115145 | 0.618594 | -0.17735 | 0.050305 |
| Staffing Organizational Units | -0.81109 | -0.09717 | 0.145557 | -0.04691 | -0.13504 | -0.14628 | 0.08815 |
| Identifying Objects Actions and Events | -0.68021 | -0.10092 | 0.09819 | -0.02084 | 0.467896 | 0.185994 | -0.01655 |
| Importance of Being Exact or Accurate | -0.01613 | -0.10875 | -0.04368 | -0.63292 | 0.41302 | 0.109074 | 0.114767 |
| Coaching and Developing Others | -0.8598 | -0.12059 | 0.197402 | 0.09274 | -0.02996 | -0.10631 | 0.008364 |
| Communicating with Supervisors Peers or Subordinates | -0.83819 | -0.12957 | 0.120982 | -0.09482 | 0.027819 | 0.184918 | -0.12529 |
| Resolving Conflicts and Negotiating with Others | -0.69509 | -0.13244 | 0.467915 | -0.00152 | -0.15645 | 0.056345 | 0.128229 |
| Developing Objectives and Strategies | -0.87997 | -0.14276 | -0.0221 | 0.127332 | 0.005041 | 0.096677 | 0.103237 |
| Contact With Others | -0.12695 | -0.16212 | 0.745163 | -0.13234 | -0.02464 | 0.004271 | 0.140374 |
| Performing for or Working Directly with the Public | -0.18409 | -0.16673 | 0.616694 | 0.31297 | 0.07541 | -0.02697 | 0.362188 |
| Provide Consultation and Advice to Others | -0.88432 | -0.17499 | -0.01272 | 0.004741 | 0.004344 | 0.061285 | 0.104229 |
| Documenting Recording Information | -0.66593 | -0.18051 | 0.10345 | -0.0959 | 0.418249 | 0.364211 | -0.02823 |
| Thinking Creatively | -0.76005 | -0.18179 | -0.2417 | 0.14907 | 0.034031 | 0.068389 | 0.239848 |
| Selling or Influencing Others | -0.53817 | -0.18751 | 0.115495 | 0.083644 | -0.29077 | -0.03541 | 0.437924 |
| Telephone | -0.40731 | -0.20397 | 0.364571 | -0.17189 | -0.0326 | 0.391764 | 0.469409 |
| Updating and Using Relevant Knowledge | -0.74367 | -0.20584 | -0.04191 | -0.03217 | 0.358433 | 0.286365 | 0.159798 |
| Deal With External Customers | -0.10282 | -0.21464 | 0.690285 | 0.011145 | -0.00516 | 0.103903 | 0.423422 |
| Organizing Planning and Prioritizing Work | -0.80767 | -0.22504 | 0.039594 | -0.01458 | -0.02134 | 0.1628 | 0.195528 |
| Structured versus Unstructured Work | -0.42256 | -0.22626 | 0.067255 | -0.00152 | 0.030399 | 0.157019 | 0.595834 |
| Public Speaking | -0.55989 | -0.23473 | 0.182039 | 0.23829 | -0.19024 | 0.060919 | 0.054316 |
| Analyzing Data or Information | -0.7818 | -0.24765 | -0.12866 | -0.06564 | 0.231512 | 0.346704 | 0.030342 |
| Processing Information | -0.73059 | -0.25275 | -0.10241 | -0.15225 | 0.299141 | 0.364526 | -0.0315 |
| Interpreting the Meaning of Information for Others | -0.79137 | -0.26497 | -0.05911 | 0.081746 | 0.199585 | 0.259669 | -0.0118 |
| Letters and Memos | -0.50735 | -0.27472 | 0.360844 | -0.1381 | -0.06266 | 0.380159 | 0.244926 |
| Communicating with Persons Outside Organization | -0.67468 | -0.28562 | 0.197973 | 0.144528 | -0.10905 | 0.288031 | 0.350471 |
| Performing Administrative Activities | -0.687 | -0.29146 | 0.224355 | -0.0265 | -0.01983 | 0.209303 | 0.203049 |
| Getting Information | -0.72961 | -0.3314 | 0.054263 | 0.007698 | 0.191659 | 0.350375 | 0.098307 |
| Establishing and Maintaining Interpersonal Relationships | -0.65022 | -0.34074 | 0.363493 | 0.097453 | -0.05525 | 0.139804 | 0.185082 |
| Interacting With Computers | -0.57543 | -0.3705 | -0.12428 | -0.24941 | 0.151814 | 0.35843 | 0.045019 |
| Electronic Mail | -0.57873 | -0.41847 | 0.084405 | -0.12674 | 0.000927 | 0.431295 | 0.251776 |
| Spend Time Sitting | -0.3079 | -0.51282 | -0.0863 | -0.19699 | -0.09264 | 0.624883 | 0.118858 |
| Indoors Environmentally Controlled | -0.27165 | -0.67834 | 0.103572 | -0.16677 | 0.200639 | 0.059161 | 0.131788 |

**Supplemental Table 2.** Factor Loadings with Varimax Rotation for O*NET Worker Characteristics

|  | Reasoning Ability | Visual-perceptual Ability | Social Ability | Coordination Skills | Creativity |
| --- | --- | --- | --- | --- | --- |
| Achievement Effort styles | -0.5800697 | 0.2695542 | 0.35567982 | 0.02236385 | -0.3357884 |
| Adaptability Flexibility styles | -0.3697493 | 0.12029496 | 0.71460848 | 0.00249894 | -0.1166679 |
| Analytical Thinking styles | -0.7889418 | 0.23809555 | 0.12461898 | 0.03584171 | -0.1289251 |
| Attention to Detail styles | -0.3979113 | 0.27298555 | 0.27548445 | 0.43079605 | 0.08747009 |
| Concern for Others styles | 0.04108898 | -0.0360425 | 0.86899306 | -0.0167346 | -0.0388367 |
| Cooperation styles | -0.138055 | 0.14522454 | 0.79238743 | 0.00400923 | 0.04384367 |
| Dependability styles | -0.2203362 | 0.06458387 | 0.75554034 | 0.06976783 | 0.00365986 |
| Independence styles | -0.2556469 | 0.19422183 | 0.45490843 | 0.02834644 | -0.3332492 |
| Initiative styles | -0.5896483 | 0.18505886 | 0.44538172 | -0.0593007 | -0.331191 |
| Innovation styles | -0.4884377 | 0.1499806 | 0.19332072 | 0.07186386 | -0.5649902 |
| Integrity styles | -0.4340342 | 0.30736307 | 0.58342426 | -0.1155212 | 0.04679538 |
| Leadership styles | -0.4629617 | -0.1007514 | 0.5857497 | -0.1997682 | -0.1422684 |
| Persistence styles | -0.6006495 | 0.18400243 | 0.38488552 | -0.0144805 | -0.3093696 |
| Self Control styles | 0.00195359 | -0.0270555 | 0.88691473 | -0.0949458 | 0.08180478 |
| Social Orientation styles | 0.06972818 | 0.02901767 | 0.86330182 | -0.133636 | -0.0655816 |
| Stress Tolerance styles | -0.2338635 | 0.07056699 | 0.79604767 | -0.0247275 | 0.12159266 |
| Arm Hand Steadiness abilities | 0.23491011 | -0.5344893 | -0.0785971 | 0.70882098 | -0.0002126 |
| Auditory Attention abilities | -0.0101648 | -0.7016074 | -0.03103 | 0.29420533 | 0.26112462 |
| Category Flexibility abilities | -0.8549898 | 0.20999761 | -0.045184 | 0.08456121 | -0.0555224 |
| Control Precision abilities | 0.18419753 | -0.6451899 | -0.2040789 | 0.5867813 | 0.10412594 |
| Deductive Reasoning abilities | -0.9013502 | 0.22524957 | 0.11925295 | -0.1134382 | -0.016926 |
| Depth Perception abilities | -0.0067199 | -0.791814 | -0.2596136 | 0.35510844 | 0.06570616 |
| Dynamic Flexibility abilities | 0.3294392 | -0.4172401 | -0.0434405 | 0.05648275 | -0.3160095 |
| Dynamic Strength abilities | 0.42266898 | -0.7757659 | -0.029102 | 0.2467387 | -0.1148348 |
| Explosive Strength abilities | 0.21021075 | -0.5617683 | 0.12867051 | 0.03689213 | -0.0652039 |
| Extent Flexibility abilities | 0.43307092 | -0.7536722 | -0.0596515 | 0.31057564 | -0.0765376 |
| Far Vision abilities | -0.5289845 | -0.537692 | -0.008805 | 0.01125217 | 0.09038468 |
| Finger Dexterity abilities | 0.06059085 | -0.3855227 | -0.1202115 | 0.78913176 | 0.07972447 |
| Flexibility of Closure abilities | -0.7841656 | -0.1970115 | -0.0184859 | 0.25731313 | 0.20296655 |
| Fluency of Ideas abilities | -0.8484202 | 0.22087216 | 0.13193085 | -0.1286725 | -0.2325811 |
| Glare Sensitivity abilities | 0.11599723 | -0.8652961 | -0.2002132 | -0.065194 | 0.11166673 |
| Gross Body Coordination abilities | 0.41238967 | -0.7953451 | 0.06344764 | 0.19028824 | -0.0917713 |
| Gross Body Equilibrium abilities | 0.31591462 | -0.8273849 | 0.02263571 | 0.1478954 | -0.0772369 |
| Hearing Sensitivity abilities | -0.1201031 | -0.6428383 | -0.0446253 | 0.38277293 | 0.16256519 |
| Inductive Reasoning abilities | -0.8763838 | 0.19160955 | 0.15792883 | -0.0489732 | -0.0406527 |
| Information Ordering abilities | -0.8966362 | 0.13000255 | 0.03101665 | 0.09641808 | 0.02034913 |
| Manual Dexterity abilities | 0.30366548 | -0.5853008 | -0.1150588 | 0.64779577 | 0.01913109 |
| Mathematical Reasoning abilities | -0.8068976 | 0.25801507 | -0.138693 | -0.0434804 | 0.14382833 |
| Memorization abilities | -0.7387354 | 0.06294183 | 0.26327519 | -0.1621511 | 0.01280279 |
| Multi-limb Coordination abilities | 0.30242189 | -0.7927145 | -0.0907513 | 0.40618741 | -0.0196016 |
| Near Vision abilities | -0.6383906 | 0.28326131 | 0.03428833 | 0.29799255 | 0.11769861 |
| Night Vision abilities | 0.08402042 | -0.8552736 | -0.1849915 | -0.1211294 | 0.14394905 |
| Number Facility abilities | -0.7603749 | 0.22254395 | -0.1200692 | -0.0388023 | 0.21878986 |
| Oral Comprehension abilities | -0.8183578 | 0.29864343 | 0.20037964 | -0.1567816 | -0.0978459 |
| Oral Expression abilities | -0.7785978 | 0.3246096 | 0.25127077 | -0.2522207 | -0.1023021 |
| Originality abilities | -0.8253482 | 0.20048993 | 0.12216382 | -0.1199447 | -0.3063454 |
| Perceptual Speed abilities | -0.5192087 | -0.4169598 | -0.0760229 | 0.37657495 | 0.40674008 |
| Peripheral Vision abilities | 0.12520063 | -0.8852389 | -0.176581 | -0.1374734 | 0.11683461 |
| Problem Sensitivity abilities | -0.838544 | 0.04230721 | 0.27183631 | 0.01754581 | 0.06992072 |
| Rate Control abilities | 0.22757705 | -0.7779328 | -0.2491206 | 0.31935008 | 0.12724719 |
| Reaction Time abilities | 0.21231299 | -0.8123556 | -0.1740063 | 0.32551056 | 0.14378708 |
| Response Orientation abilities | 0.17665116 | -0.8431334 | -0.1011003 | 0.28397444 | 0.13028738 |
| Selective Attention abilities | -0.5151358 | -0.3727775 | 0.03467998 | 0.23576711 | 0.25329178 |
| Sound Localization abilities | 0.09716875 | -0.8557227 | -0.166117 | -0.0551106 | 0.14129215 |
| Spatial Orientation abilities | 0.06971622 | -0.870398 | -0.2101497 | -0.1183657 | 0.10786482 |
| Speech Clarity abilities | -0.5983201 | 0.25510392 | 0.37278161 | -0.4071955 | -0.1739028 |
| Speech Recognition abilities | -0.5898991 | 0.30858031 | 0.43924066 | -0.2910597 | 0.07511563 |
| Speed of Closure abilities | -0.807733 | -0.1280125 | 0.17898219 | 0.04777447 | 0.17843939 |
| Speed of Limb Movement abilities | 0.36620572 | -0.8070975 | -0.0515014 | 0.11855033 | 0.00853607 |
| Stamina abilities | 0.44501608 | -0.7756135 | 0.06407869 | 0.19667986 | -0.0905654 |
| Static Strength abilities | 0.4198236 | -0.7841333 | 0.00106003 | 0.29722767 | -0.0700701 |
| Time Sharing abilities | -0.4119565 | -0.5151815 | 0.29674498 | -0.0526184 | 0.23330987 |
| Trunk Strength abilities | 0.44079937 | -0.7208807 | 0.02874232 | 0.23431235 | -0.1476171 |
| Visual Color Discrimination abilities | -0.2644554 | -0.4625094 | -0.1840066 | 0.5974126 | 0.04581152 |
| Visualization abilities | -0.4843938 | -0.4143914 | -0.2795844 | 0.46419702 | -0.1205811 |
| Wrist Finger Speed abilities | 0.22553687 | -0.539262 | -0.2336406 | 0.53242234 | 0.08627292 |
| Written Comprehension abilities | -0.8265127 | 0.38688427 | 0.14008552 | -0.1614834 | -0.0640422 |
| Written Expression abilities | -0.7968129 | 0.38685274 | 0.19054717 | -0.2359784 | -0.0944337 |
| Artistic interests | -0.2467304 | 0.27487506 | 0.09220041 | -0.0448751 | -0.6505481 |
| Conventional interests | 0.14221114 | 0.27930511 | -0.0425875 | -0.0882116 | 0.65242672 |
| Enterprising interests | -0.1131237 | 0.19122605 | 0.29911177 | -0.5165951 | 0.19489632 |
| Investigative interests | -0.703497 | 0.15061978 | -0.1338746 | 0.18002389 | -0.1148319 |
| Realistic interests | 0.22951131 | -0.5862171 | -0.3994587 | 0.49553296 | 0.04907839 |
| Social interests | -0.0900165 | 0.20204009 | 0.70781932 | -0.2577675 | -0.2447503 |
| Achievement values | -0.7906933 | 0.23794573 | 0.26955333 | -0.1421547 | -0.276233 |
| Independence values | -0.7448816 | 0.1088891 | 0.26305126 | -0.2133973 | -0.1918647 |
| Recognition values | -0.8160306 | 0.24667829 | 0.23171567 | -0.1594597 | -0.1855496 |
| Relationships values | -0.0686302 | 0.10324931 | 0.7317915 | -0.2149127 | -0.0519426 |
| Support values | -0.0965588 | -0.3265627 | 0.10598608 | 0.27233887 | 0.56256385 |
| Working Conditions values | -0.8328684 | 0.18194311 | 0.20996792 | -0.1415398 | -0.151944 |

**Supplemental Table 3.** Factor Loadings with Varimax Rotation for O*NET Worker Requirements

|  | Analytical Skills | Technical Skills | Health Services Knowledge | Business Skills | Humanities Knowledge |
| --- | --- | --- | --- | --- | --- |
| Active Learning skills | -0.8598502 | 0.11144623 | -0.318518 | 0.19507911 | -0.1253035 |
| Active Listening skills | -0.7446759 | 0.31968558 | -0.3728173 | 0.21494632 | -0.1649436 |
| Complex Problem Solving skills | -0.8618688 | -0.0358362 | -0.2594827 | 0.23202091 | -0.1144741 |
| Coordination skills | -0.4707837 | -0.0036941 | -0.412473 | 0.54445691 | -0.0535257 |
| Critical Thinking skills | -0.8288778 | 0.13886661 | -0.3063707 | 0.25077959 | -0.1281976 |
| Equipment Maintenance skills | 0.20532329 | -0.7707925 | 0.09287306 | -0.1937889 | 0.21683017 |
| Equipment Selection skills | 0.00430705 | -0.797125 | 0.10573289 | -0.1988554 | 0.25313781 |
| Installation skills | -0.0500384 | -0.555332 | 0.17669068 | -0.1207008 | 0.11390908 |
| Instructing skills | -0.7041556 | 0.0404982 | -0.462866 | 0.18805304 | -0.1272221 |
| Judgment and Decision Making skills | -0.8223607 | 0.06460565 | -0.309854 | 0.29527768 | -0.1063056 |
| Learning Strategies skills | -0.7175634 | 0.10229056 | -0.4506681 | 0.17662763 | -0.1449135 |
| Management of Financial Resources skills | -0.4835369 | -0.0724009 | 0.03881 | 0.69270627 | 0.09067939 |
| Management of Material Resources skills | -0.4668144 | -0.2484608 | -0.0645712 | 0.62268182 | 0.05743321 |
| Management of Personnel Resources skills | -0.5656086 | -0.0698543 | -0.35325 | 0.5542023 | -0.0354564 |
| Mathematics skills | -0.8037376 | -0.1657662 | 0.0902339 | 0.18367075 | 0.01645094 |
| Monitoring skills | -0.7206784 | -0.0091733 | -0.4298296 | 0.31676042 | -0.0701724 |
| Negotiation skills | -0.5016186 | 0.25739595 | -0.2898421 | 0.61302301 | -0.0667234 |
| Operation and Control skills | 0.26479607 | -0.8166213 | -0.013625 | -0.0800481 | 0.20381221 |
| Operation Monitoring skills | 0.05109343 | -0.8543488 | -0.0254481 | -0.0486645 | 0.26579669 |
| Operations Analysis skills | -0.7273366 | -0.1143787 | -0.0193248 | 0.1739208 | -0.1141595 |
| Persuasion skills | -0.5878922 | 0.25724146 | -0.3251738 | 0.54533003 | -0.0668278 |
| Programming skills | -0.7861536 | -0.0893678 | 0.25089475 | -0.0344519 | 0.04546454 |
| Quality Control Analysis skills | -0.1165029 | -0.8427935 | 0.03195039 | -0.0397804 | 0.24903426 |
| Reading Comprehension skills | -0.8572325 | 0.18766884 | -0.3051538 | 0.10989 | -0.1795213 |
| Repairing skills | 0.19267805 | -0.7697531 | 0.10735642 | -0.1776125 | 0.21029901 |
| Science skills | -0.7153792 | -0.2755537 | -0.3601307 | -0.1842422 | -0.0957513 |
| Service Orientation skills | -0.3610433 | 0.37341456 | -0.5158855 | 0.41976058 | 0.00080391 |
| Social Perceptiveness skills | -0.4667515 | 0.34183816 | -0.6204089 | 0.35596537 | -0.0793518 |
| Speaking skills | -0.7437979 | 0.32058148 | -0.3501402 | 0.24438576 | -0.2319586 |
| Systems Analysis skills | -0.8454341 | -0.0467414 | -0.1737963 | 0.32845711 | -0.0821722 |
| Systems Evaluation skills | -0.834801 | -0.0514102 | -0.2291927 | 0.34468833 | -0.0679766 |
| Technology Design skills | -0.647987 | -0.4175904 | 0.15050785 | 0.03472582 | 0.06973939 |
| Time Management skills | -0.6566232 | 0.03825024 | -0.3305597 | 0.45761861 | -0.0601244 |
| Troubleshooting skills | 0.07054643 | -0.882313 | 0.06286483 | -0.1292857 | 0.24159037 |
| Writing skills | -0.8176031 | 0.25977543 | -0.2763621 | 0.1556708 | -0.2350147 |
| Administration and Management knowledge | -0.4801473 | -0.028396 | -0.0416528 | 0.68756321 | -0.2266427 |
| Biology knowledge | -0.3747383 | -0.1692043 | -0.5905816 | -0.0638801 | -0.1514011 |
| Building and Construction knowledge | 0.0314458 | -0.6683665 | 0.17647549 | 0.25650249 | -0.250258 |
| Chemistry knowledge | -0.2628511 | -0.6447964 | -0.3202476 | -0.0677868 | -0.0637445 |
| Clerical knowledge | -0.4206749 | 0.34269072 | -0.1109993 | 0.42140048 | -0.2063467 |
| Communications and Media knowledge | -0.4668172 | 0.33278075 | -0.1324225 | 0.25275534 | -0.5320559 |
| Computers and Electronics knowledge | -0.7788493 | 0.01868908 | 0.07793847 | 0.12891216 | -0.2181142 |
| Customer and Personal Service knowledge | -0.1000085 | 0.26709703 | -0.3908816 | 0.54866202 | -0.0999962 |
| Design knowledge | -0.3737125 | -0.6377951 | 0.35524409 | 0.12517579 | -0.2652131 |
| Economics and Accounting knowledge | -0.3364577 | 0.17950055 | 0.11978383 | 0.66224184 | -0.1116788 |
| Education and Training knowledge | -0.4696746 | -0.0002036 | -0.5076284 | 0.18170388 | -0.411361 |
| Engineering and Technology knowledge | -0.4511066 | -0.7503641 | 0.2639456 | 0.07331324 | -0.1571143 |
| English Language knowledge | -0.6502253 | 0.31537336 | -0.2746551 | 0.14111086 | -0.4431289 |
| Fine Arts knowledge | -0.0166553 | 0.23370246 | 0.03835948 | 0.02759682 | -0.4721215 |
| Food Production knowledge | 0.16772199 | -0.1482077 | -0.0784375 | 0.22739163 | -0.1345968 |
| Foreign Language knowledge | -0.1175628 | 0.1389325 | -0.341774 | 0.07545146 | -0.5835189 |
| Geography knowledge | -0.2894827 | -0.0312704 | -0.0201771 | 0.22503813 | -0.7172398 |
| History and Archeology knowledge | -0.2399929 | 0.21414265 | -0.186409 | 0.02198083 | -0.7870117 |
| Law and Government knowledge | -0.3398426 | 0.07595149 | -0.3106572 | 0.43260686 | -0.403659 |
| Mathematics knowledge | -0.7463478 | -0.2185493 | 0.13795868 | 0.16982605 | -0.1488835 |
| Mechanical knowledge | 0.03962823 | -0.9164118 | 0.12058387 | 0.01095254 | 0.01025931 |
| Medicine and Dentistry knowledge | -0.1416078 | -0.0058122 | -0.8023338 | -0.0592466 | 0.08379672 |
| Personnel and Human Resources knowledge | -0.4059916 | 0.04707988 | -0.3046751 | 0.64866362 | -0.2311557 |
| Philosophy and Theology knowledge | -0.1769729 | 0.36849903 | -0.6063036 | 0.05864152 | -0.4945659 |
| Physics knowledge | -0.415584 | -0.7474777 | -0.026726 | -0.0746062 | -0.1647695 |
| Production and Processing knowledge | -0.0229729 | -0.6170339 | 0.30025353 | 0.26159484 | 0.05085698 |
| Psychology knowledge | -0.1992297 | 0.25471199 | -0.7775439 | 0.23137883 | -0.2122373 |
| Public Safety and Security knowledge | 0.09603459 | -0.5213617 | -0.3432535 | 0.34794979 | -0.2911874 |
| Sales and Marketing knowledge | -0.2087133 | 0.19412581 | 0.1445255 | 0.60361099 | -0.1267942 |
| Sociology and Anthropology knowledge | -0.2654915 | 0.3909558 | -0.56939 | 0.12526567 | -0.4933873 |
| Telecommunications knowledge | -0.3310745 | -0.2698527 | 0.04380014 | 0.26367433 | -0.2758193 |
| Therapy and Counseling knowledge | -0.1084052 | 0.24745941 | -0.844979 | 0.09590907 | -0.1052651 |
| Transportation knowledge | 0.19855136 | -0.4491931 | 0.00100023 | 0.41976422 | -0.3513124 |

**Supplemental Table 4.** Multiple linear regression models of cognitive dysfunction with O*NET Occupation Requirements (A), Worker Characteristics (B), and Worker Requirements (C) covarying for C9 expansion status in a subset of the overall sample with available genetic data

| **A. Occupational Requirements** | **ECAS Total Score** | **ECAS ALS Specific Score** | **ECAS ALS Non-Specific Score** |
| --- | --- | --- | --- |
| Model R^2^ | 0.28 | 0.23 | 0.16 |
| F-statistic | 4.33 | 3.58 | 3.43 |
| DF | 11, 85 | 11, 85 | 11, 85 |
| *p* | 3.92e-06 | .0003 | .015 |
|  | β (S.E.) | β (S.E.) | β (S.E.) |
| Intercept | 110.70 (8.15)*** | 83.69 (7.27)*** | 27.01 (3.41)*** |
| Age at Test | -0.25 (0.09)** | -0.18 (0.08)* | -0.07 (0.04) |
| Education | 0.88 (0.40)* | 0.56 (0.36) | 0.31 (0.17) |
| Non-Bulbar Onset | 0.23 (2.22) | 0.13 (1.98) | 0.11 (0.93) |
| C9 Expansion Positive | -2.09 (2.98) | -1.33 (2.67) | -0.77 (1.24) |
| Managerial Capacity | -1.85 (1.25) | -1.99 (1.12) | 0.15 (0.52) |
| Exposures to Job Hazards | -3.38 (1.12)** | -3.16 (1.00) | -0.19 (0.47) |
| Exposures to Conflict | 0.25 (1.01) | -0.68 (0.90) | 0.93 (.042)* |
| Precision Skills | 0.64 (0.95) | 0.64 (0.85) | 0.01 (0.39) |
| Healthcare | -0.31 (0.91) | -0.02 (0.81) | -0.29 (0.38) |
| Physical Ability | 0.84 (1.01) | 0.58 (0.90) | 0.27 (0.42) |
| Autonomy | 1.57 (1.09) | 1.34 (0.98) | 0.22 (0.45) |
| **B. Worker Characteristics** | **ECAS Total Score** | **ECAS ALS Specific Score** | **ECAS ALS Non-Specific Score** |
| Model R^2^ | 0.34 | 0.21 | 0.08 |
| F-statistic | 5.06 | 3.87 | 1.87 |
| DF | 9, 87 | 9, 87 | 9, 87 |
| *P* | 1.27e-06 | .0003 | .075 |
|  | β (S.E.) | β (S.E.) | β (S.E.) |
| Intercept | 112.00 (8.24)*** | 85.38 (7.43)*** | 26.62 (3.40)*** |
| Age at Test | -0.28 (0.08)** | -0.21 (0.08)* | -0.07 (0.04) |
| Education | 0.93 (0.40)* | 0.59 (0.36) | 0.34 (0.17)* |
| Non-Bulbar Onset | -0.20 (2.21) | -0.29 (1.99) | 0.09 (0.91) |
| C9 Expansion Positive | -1.88 (2.92) | -0.97 (2.63) | -0.92 (1.21) |
| Reasoning Ability | -1.72 (1.13) | -1.85 (1.01) | 0.14 (0.46) |
| Visual Perceptual Ability | 1.82 (1.00) | 1.95 (0.90)* | -0.13 (0.41) |
| Social Ability | 1.84 (0.87)* | 1.10 (0.78) | 0.74 (0.36)* |
| Coordination Skills | -1.02 (0.85) | -0.51 (0.77) | -0.52 (0.35) |
| Creativity | -0.69 (1.11) | -0.91 (1.00) | 0.21 (0.46) |
| **C. Worker Requirements** | **ECAS Total Score** | **ECAS ALS Specific Score** | **ECAS ALS Non-Specific Score** |
| Model R^2^ | 0.31 | 0.23 | 0.10 |
| F-statistic | 5.70 | 4.25 | 2.12 |
| DF | 9, 87 | 9, 87 | 9, 87 |
| *p* | 3.45e-07 | .0001 | .095 |
|  | β (S.E.) | β (S.E.) | β (S.E.) |
| Intercept | 112.31 (8.06)*** | 85.20 (7.32)*** | 27.11 (3.36)*** |
| Age at Test | -0.27 (0.09)** | -0.20 (0.07)* | -0.06 (0.04) |
| Education | 0.87 (0.39)* | 0.58 (0.35) | 0.29 (0.16) |
| Non-Bulbar Onset | 0.18 (2.15) | -0.13 (1.96) | 0.31 (0.90) |
| C9 Expansion Positive | -1.54 (2.88) | -0.83 (2.61) | -0.71 (1.19) |
| Analytical Skills | -2.69 (1.25)* | -2.82 (1.14)* | 0.14 (0.52) |
| Technical Skills | 2.17 (0.89)* | 1.69 (0.81)* | 0.48 (0.37) |
| Health Services Knowledge | -0.35 (0.91) | -0.15 (0.83) | -0.20 (0.38) |
| Business skills/Knowledge | 0.95 (0.66) | 0.42 (0.60) | 0.53 (0.27) |
| Humanities Knowledge | -2.32 (0.86)** | -1.59 (0.78)* | -0.72 (0.36)* |

**Supplemental Table 5.** Multiple linear regression models of motor dysfunction with O*NET Occupation Requirements (A), Worker Characteristics (B), and Worker Requirements (C) covarying for C9 expansion status in a subset of the overall sample with available genetic data

| **A. Occupational Requirements** | **UMN Total Score** | **ALSFRS-R Total Score** |
| --- | --- | --- |
| Model R^2^ | 0.19 | 0.03 |
| F-statistic | 2.86 | 1.23 |
| DF | 11, 85 | 11, 85 |
| *p* | .003 | .282 |
|  | β (S.E.) | β (S.E.) |
| Intercept | 30.18 (8.15)*** | 41.25 (6.55)*** |
| Age at Test | -0.17 (0.09) | -0.04 (0.07) |
| Education | -0.93 (0.40)* | 0.04 (0.31) |
| Non-Bulbar Onset | 2.35 (2.13) | -5.54 (1.87)** |
| C9 Expansion Status | 1.18 (2.91) | -0.80 (2.33) |
| Managerial Capacity | -2.64 (1.23)* | 0.09 (1.06) |
| Exposures to job hazards | -2.36 (1.09)* | -0.11 (0.95) |
| Exposures to conflict | 0.47 (0.99) | -1.20 (0.89) |
| Precision skills | -2.05 (0.90)* | 0.54 (0.83) |
| Healthcare | 1.85 (0.87)* | -0.32 (0.78) |
| Physical ability | -0.36 (0.94) | 0.08 (0.92) |
| Autonomy | 1.89 (1.08) | 0.21 (0.96) |
| **B. Worker Characteristics** | **UMN Total Score** | **ALSFRS-R Total Score** |
| Model R^2^ | 0.08 | 0.05 |
| F-statistic | 1.78 | 1.54 |
| DF | 9, 87 | 9, 87 |
| *p* | .085 | .147 |
|  | β (S.E.) | β (S.E.) |
| Intercept | 29.67 (8.79)** | 41.32 (6.43)*** |
| Age at Test | -0.21 (0.09)* | -0.05 (0.07) |
| Education | -0.67 (0.42) | 0.06 (0.30) |
| Non-Bulbar Onset | 1.75 (2.25) | -5.36 (1.82)** |
| C9 Expansion Status | 0.86 (3.04) | -1.13 (2.27) |
| Reasoning Ability | -2.51 (1.18)* | 0.23 (0.87) |
| Visual-perceptual Ability | 2.07 (0.98)* | -0.01 (0.81) |
| Social ability | 0.78 (0.91) | -1.06 (0.70) |
| Coordination skills | 1.78 (0.86)* | -0.22 (0.66) |
| Creativity | -0.11 (1.14) | 0.73 (0.90) |
| **C. Worker Requirements** | **UMN Total Score** | **ALSFRS-R Total Score** |
| Model R^2^ | 0.01 | 0.07 |
| F-statistic | 1.05 | 1.80 |
| DF | 9, 87 | 9, 87 |
| *p* | .412 | .080 |
|  | β (S.E.) | β (S.E.) |
| Intercept | 21.66 (8.82)* | 41.12 (6.42)*** |
| Age at Test | -0.16 (0.09) | -0.04 (0.06) |
| Education | -0.33 (0.42) | 0.06 (0.30) |
| Non-Bulbar Onset | 1.40 (2.35) | -5.58 (1.80)** |
| C9 Expansion Status | 1.05 (3.15) | -0.76 (2.25) |
| Analytical skills | -1.98 (1.41) | -0.16 (0.99) |
| Technical skills | 0.99 (0.97) | -0.58 (0.71) |
| Health services knowledge | -1.24 (1.02) | 1.54 (0.73)* |
| Business skill | 0.20 (0.72) | -0.22 (0.50) |
| Humanities knowledge | 0.35 (0.95) | 0.50 (0.68) |
